# Post-Concussion Assessment as a diagnostic and mechanistic framework for treating patients with Long COVID

**DOI:** 10.1101/2022.09.24.22280310

**Authors:** Bradley S. Davidson, Lily Noteboom, Hannah Pierro, Cayce Kantor, Daniel Stoot, Fred Stoot, Daniel Linseman, Troy Hale, Kim Gorgens

## Abstract

**Introduction:** Despite first coming into view over two years ago, effective diagnostic and treatment pathways for Long COVID continue to evade the medical community. The overlap in neurological-based symptoms and neuroinflammatory origin indicates that the framework of post-concussion syndrome may provide insight into new diagnostics and treatment for patients with Long COVID. The objective of this investigation was to determine whether tools from the four common domains of concussion assessment were sensitive to differentiate between patients with Long COVID from a reference group who was infected with Sars-CoV-2 and does not have Long COVID.

**Methods:** In this prospective cohort design, each participant self reported their group (Acute, n=28) and Long COVID Group (n=33). Each participant underwent an examination in four assessment categories: symptoms, vestibular nystagmography, Automated Neuropsychological Assessment Metrics (ANAM), and a series of balance tasks.

**Results:** Total Symptom scores were separated into functional classifications and showed clear success as a tool to differentiate between Acute and Long COVID. Five of the 33 people in the Long COVID had detectable central lesions, which increases the risk of developing long COVID by 64% (Relative Risk=1.64). A wide variety of objective and quantitative measures from post-concussion care are sensitive to the Long COVID condition. Prolonged latency during random saccades eye tracking was present (p<0.01, d=0.87) in the Long COVID group corresponding to the King-Devick rapid reading test, which was highly sensitive to Long COVID (p<0.01, d=1.34). ANAM reaction time subtests had similarly large effects (p<0.01, d=0.93-1.09). Balance performance with corrupted sensory feedback was also sensitive (p<0.01, d=0.96).

**Discussion:** Our results indicate that long-standing and validated post-concussion symptom questionnaires may be used for quantifying the severity of Long COVID. Some of the most sensitive measures (especially the King-Devick rapid reading test) are easy to implement clinically and may be effective at tracking patient progress in the context of Long COVID treatment. The results point to wide deficits in motor integration and provide a rationale for treating the subset of Long COVID patients with similar rehabilitation strategies as patients with post-concussion syndrome.

## Introduction

Despite first coming into view over two years ago, effective diagnostic and treatment pathways for Long COVID continue to evade the medical community. Reliable diagnostic and treatment options are sparse and often unsuccessful. Some sources estimate that more than 40% of US adults have experienced COVID-19 and roughly 1 in 5 of those adults are experiencing Long COVID (CDC, 2022). From the CDC: Long COVID “…occurs in individuals with a history of probable or confirmed SARS-CoV-2 infection, usually 3 months from the onset of COVID-19 with symptoms that last for at least 2 months and cannot be explained by an alternative diagnosis…and generally have an impact on everyday functioning. Symptoms may be new onset following initial recovery from an acute COVID-19 episode or persist from the initial illness…” The effects of Long COVID appear in a variety of organ systems and a constellation of symptoms that span across multiple seemingly unrelated categories (Table 1, first column). To better understand the presentation of Long COVID, researchers and clinicians have thoroughly examined the symptom profiles. Yong and Liu (2021) performed a broad literature search and classified Long COVID into six subtypes that considered clusters of symptoms and organ systems. In parallel, Davis et al. (2021) identified three clusters (i.e. subtypes) based on onset, trajectory, and duration of symptoms over a 7 month period after COVID-19 infection. Most recently, a retrospective investigation with a large sample size verified the three-phenotype classification as patients predominantly having: 1) broad spectrum of symptoms, 2) respiratory symptoms, and 3) mental health and cognitive symptoms (Subramanian et al. 2022).

**Table 1.**
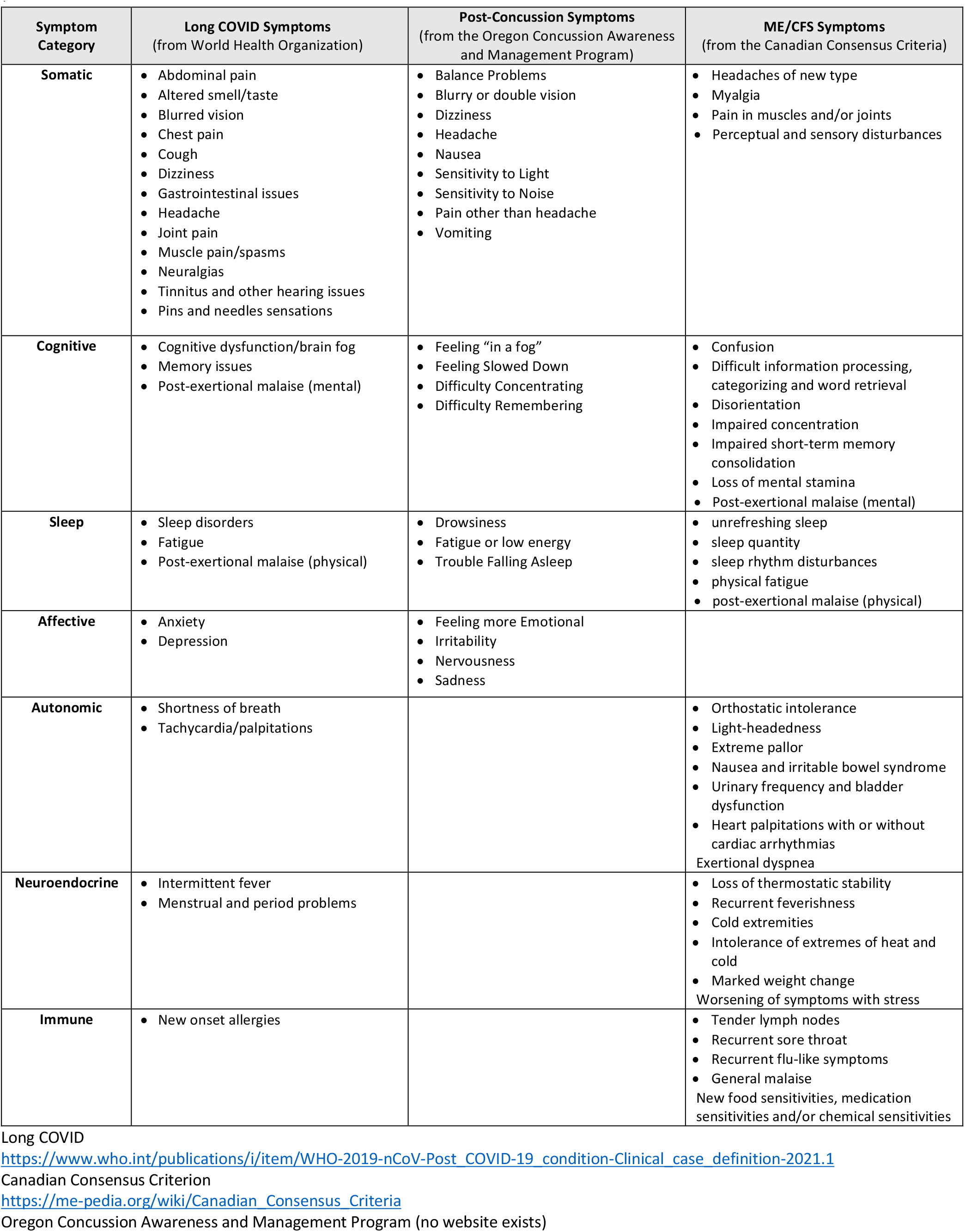
Long COVID symptoms taken from WHO clinical case definition from Oct 6, 2021. The Post-concussion Symptoms listed were taken from the post-concussion symptom questionnaire used in this investigation and mirror many available post-concussion symptom questionnaires used to assess an athlete’s status within the acute phase of a concussive injury.

| Symptom Category | Long COVID Symptoms<br>(from World Health Organization) | Post-Concussion Symptoms<br>(from the Oregon Concussion Awareness and Management Program) | ME/CFS Symptoms<br>(from the Canadian Consensus Criteria) |
| --- | --- | --- | --- |
| **Somatic** | Abdominal pain<br>Altered smell/taste<br>Blurred vision<br>Chest pain<br>Cough<br>Dizziness<br>Gastrointestinal issues<br>Headache<br>Joint pain<br>Muscle pain/spasms<br>Neuralgias<br>Tinnitus and other hearing issues<br>Pins and needles sensations | Balance Problems<br>Blurry or double vision<br>Dizziness<br>Headache<br>Nausea<br>Sensitivity to Light<br>Sensitivity to Noise<br>Pain other than headache<br>Vomiting | Headaches of new type<br>Myalgia<br>Pain in muscles and/or joints<br>Perceptual and sensory disturbances |
| **Cognitive** | Cognitive dysfunction/brain fog<br>Memory issues<br>Post-exertional malaise (mental) | Feeling “in a fog”<br>Feeling Slowed Down<br>Difficulty Concentrating<br>Difficulty Remembering | Confusion<br>Difficult information processing, categorizing and word retrieval<br>Disorientation<br>Impaired concentration<br>Impaired short-term memory consolidation<br>Loss of mental stamina<br>Post-exertional malaise (mental) |
| **Sleep** | Sleep disorders<br>Fatigue<br>Post-exertional malaise (physical) | Drowsiness<br>Fatigue or low energy<br>Trouble Falling Asleep | unrefreshing sleep<br>sleep quantity<br>sleep rhythm disturbances<br>physical fatigue<br>post-exertional malaise (physical) |
| **Affective** | Anxiety<br>Depression | Feeling more Emotional<br>Irritability<br>Nervousness<br>Sadness |  |
| **Autonomic** | Shortness of breath<br>Tachycardia/palpitations |  | Orthostatic intolerance<br>Light-headedness<br>Extreme pallor<br>Nausea and irritable bowel syndrome<br>Urinary frequency and bladder dysfunction<br>Heart palpitations with or without cardiac arrhythmias<br>Exertional dyspnea |
| **Neuroendocrine** | Intermittent fever<br>Menstrual and period problems |  | Loss of thermostatic stability<br>Recurrent feverishness<br>Cold extremities<br>Intolerance of extremes of heat and cold<br>Marked weight change<br>Worsening of symptoms with stress |
| **Immune** | New onset allergies |  | Tender lymph nodes<br>Recurrent sore throat<br>Recurrent flu-like symptoms<br>General malaise<br>New food sensitivities, medication sensitivities and/or chemical sensitivities |

Long COVID
https://www.who.int/publications/i/item/WHO-2019-nCoV-Post_COVID-19_condition-Clinical_case_definition-2021.1
Canadian Consensus Criterion
https://me-pedia.org/wiki/Canadian_Consensus_Criteria
Oregon Concussion Awareness and Management Program (no website exists)

The neurological deficits accompanying Long COVID are widely recognized; however, it is not clear what pathophysiological mechanisms—whether structural, immunological, or otherwise—are responsible for these neurological deficits. A well-done investigation shortly after the discovery of Long COVID reported that 85% of Long COVID patients had more than 4 neurologic symptoms and 53% had an abnormal neurological exam with prominent short-term memory loss and attention functions (Graham et al. 2021). Risks of various neurological elements such as cognitive deficit, dementia, and epilepsy all remained high even two years following COVID-19 infection (Taquet et el., 2021). The presence of regional and localized structural changes detected following COVID-19 infection raises the question on whether identifiable lesions may be responsible for the neurological deficits that Long COVID patients experience. Patients who had measurable changes in the frontal cortex, parahippocampal gyrus, and olfactory cortex along with significant reduction in overall brain size, also experienced a cognitive decline six months after acute infection with COVID-19 (Douaud et al. 2022).

Recently emerging data indicate that microclotting and localized neuroinflammation are complementary factors in the development of persistent Long neurological COVID (Marzoog 2022). Markers of systemic inflammation (e.g., serum-derived IL-6, IL-8, galectin-9, etc.) have been repeatedly identified in patients with Long COVID independent of presenting symptoms (Berentschot et al. 2022, Kedor et al. 2022, Li et al. 2022, Rousseau 2022). However, because the percentage of patients with these inflammatory markers is relatively small (25-50%), it fails to explain a significant fraction of the population of Long COVID patients. Recent data from Kruger et al. (2022) suggest that insoluble inflammatory molecules and other proteins may become entrapped in a series of microclots, which allows large amounts of inflammation to go undetected in serum samples typically collected from COVID patients.

Given the overlap in neurological-based symptoms and emerging overlap in neuroinflammatory origin, the framework of post-concussion syndrome may provide insight into new diagnostics and treatment for patients with Long COVID. Although Long COVID has a viral origin it is interesting that the most common symptoms provide a surprising amount of overlap with a post-concussion condition that has an obviously neurotraumatic origin. The concussive framework provides clinically efficient tools that have evolved over time. For example, Table 1 column 2 lists the symptoms examined in a concussion symptom questionnaire used frequently with concussed athletes. Despite the simplicity of the questionnaire, the questionnaire accounts for the most prevalent symptoms—brain fog, fatigue, and emotional state—reported despite various subtypes and classifications (Davis et al. 2021).

Myalgic Encephalomyelitis/Chronic Fatigue Syndrome (ME/CFS) has been proposed as a model template for Long COVID but has two limitations. The connection of symptoms between Long COVID and ME/CFS was recognized as early as 2020 and was named as one of the six a subtypes in Yong and Liu (2021). ME/CFS framework is an umbrella term that combines two conditions (ME and CFS) that were distinct as recently as 2015 (Institute of Medicine, 2015) and disagreement still exists on whether this combination should exist (Jason et al. 2016). Combining these conditions into one disorder, the potential symptoms are expansive, and mostly align with Long COVID (Table 1), but with a major exception. The ME/CFS framework does not consider the affective category within the diagnostic criteria, which is a major gap when considering Long COVID, in which affective symptoms of anxiety, nervousness, and depression are prevalent across clusters. Another limitation is that the diagnostic criteria of post-exertional malaise (PEM) lasting longer than 14 hours, a which is *the hallmark symptom in ME/CFS*, is not applicable for most Long COVID patients. A recent investigation specifically applying the ME/CFS model found that only 45% of Long COVID patients who presented with moderate to severe fatigue actually met this PEM criterion (Kedor et al., 2022).

Our central premise is that the large body of knowledge surrounding post-concussion diagnostics and treatment can be used to guide practical approach to understanding the most common presentations of Long COVID, which are neurologically oriented. Therefore, the objective of this investigation was to determine whether tools from the four common domains of concussion assessment (symptoms, neurocognitive performance, vestibular-ocular control, balance performance) were sensitive to differentiate between patients with Long COVID from a reference group who was infected with Sars-CoV-2 and does not have Long COVID. We hypothesized that: 1) Participants in the Long COVID group would demonstrate high scores on a symptom inventory specifically designed for post-concussion tracking. 2) Participants with Long COVID group would demonstrate higher incidence of central integration deficits compared to post-COVID reference population based on: a) quantitative vestibular ocular-motility assessment, b) neurocognitive motor control tests, and b) postural control during standing balance tasks. The results of this investigation point to quantitative tools that can be easily used to assess the severity of Long COVID and be added to a comprehensive Long COVID treatment paradigm to track patient progress in symptom resolution and daily function. These results also provide a rationale for treating the subset of Long COVID patients with clear neurological symptoms by similar rehabilitation strategies as patients with post-concussion syndrome.

## Methods

### Study Design

This investigation was a prospective cohort design where we recruited participants into one of two groups: Acute or Long COVID. During the screening phone call we asked: “Have you ever been diagnosed with “Long COVID” or “COVID-19 Long Hauler’s Syndrome”?”. At the start of the test, we verified this information and placed them into the Acute or Long COVID group. Inclusion criteria: Between the ages of 18 and 75; Was infected with COVID-19; First date of symptoms was more than 12 weeks prior to scheduled testing date. Exclusion criteria: Prior concussion or other head trauma within the last 10 years; Known balance problems or dizziness prior to having COVID-19; Parkinson’s Disease; Neurodegenerative disorder; History of vestibular disease; Otolithic surgery; Chronic pain; Multiple sclerosis; Fibromyalgia; Medications that affect their cognitive ability or balance; or Use recreational drugs more than once a week. The investigation was approved by the University of Denver IRB (protocol # 1848486).

Each participant visited one of two testing sites for a single session lasting 1.5 hours, which included four assessment categories: symptoms, vestibular nystagmography, Automated Neuropsychological Assessment Metrics (ANAM), and a series of balance tasks.

### Post-COVID Symptom Assessment

The primary tools we used to describe the long-term effects of COVID in each participant were: 1) self-reported status as having Long COVID and 2) the answers to a post-concussion symptom questionnaire. The post-concussion symptom questionnaire we chose was a modified list from the Oregon Concussion Awareness and Management Program (OCAMP). This tool has a list of 22 symptoms common during the acute post-concussion status (Table 1). The participant indicated the severity of each symptom on a 7-point Likert scale that ranges from 0 to 6. The participants was prompted to “…circle the number that matches the severity. Mark a 0 is you did not have the symptom…” and the numbers were associated with the categories of None, Mild, Moderate, and Severe (Table 2). This particular post-concussion symptom questionnaire is formatted similarly to the Symptom Evaluation section from the SCAT-5 (Echemendia et al. 2017) and Post-Concussion Symptom Inventory (Gioia et al., 2008). The numbers circled were added together to create a Symptom Score, which had a possible range from 0 (no symptoms present) to 132 (all symptoms present at the most severe level).

**Table 2:** Likert Scale score that the participant assigned for a list of 22 symptoms common to post-concussion.

| Severity of Symptom | Severity Prompt |
| --- | --- |
| 0 | None |
| 1-2 | Mild |
| 3-4 | Moderate |
| 5-6 | Severe |

**Table 3:**
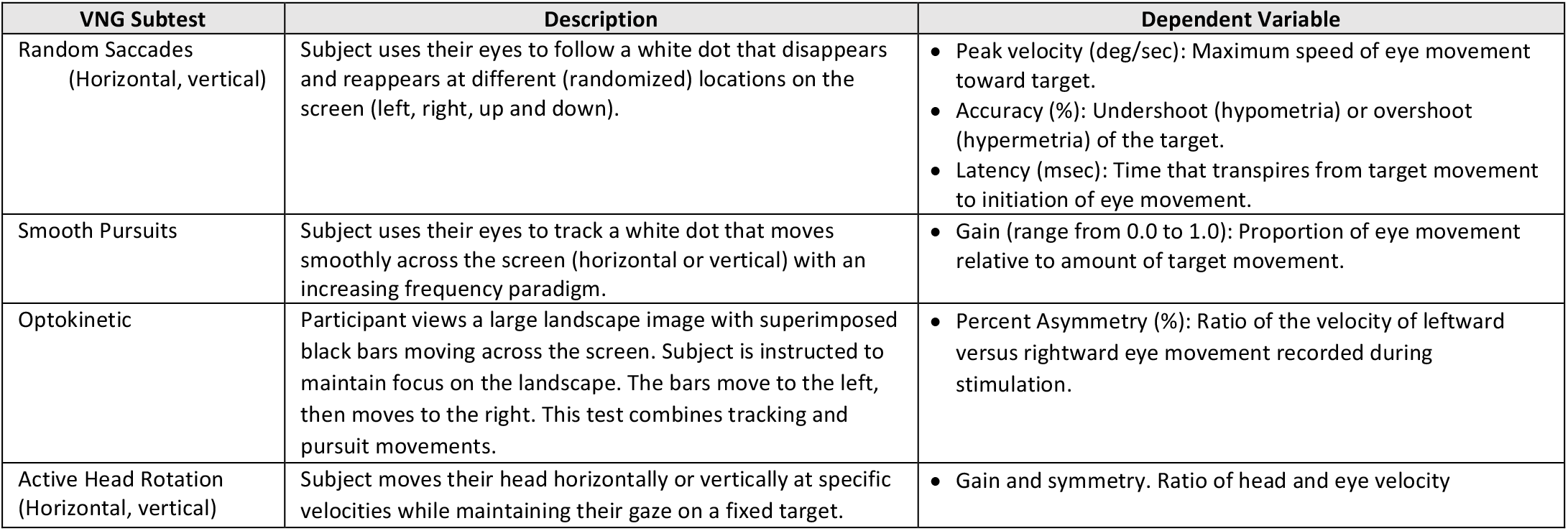
Videonystagmography subtest summary

| VNG Subtest | Description | Dependent Variable |
| --- | --- | --- |
| Random Saccades<br>(Horizontal, vertical) | Subject uses their eyes to follow a white dot that disappears and reappears at different (randomized) locations on the screen (left, right, up and down). | • Peak velocity (deg/sec): Maximum speed of eye movement toward target.<br>• Accuracy (%): Undershoot (hypometria) or overshoot (hypermetria) of the target.<br>• Latency (msec): Time that transpires from target movement to initiation of eye movement. |
| Smooth Pursuits | Subject uses their eyes to track a white dot that moves smoothly across the screen (horizontal or vertical) with an increasing frequency paradigm. | • Gain (range from 0.0 to 1.0): Proportion of eye movement relative to amount of target movement. |
| Optokinetic | Participant views a large landscape image with superimposed black bars moving across the screen. Subject is instructed to maintain focus on the landscape. The bars move to the left, then moves to the right. This test combines tracking and pursuit movements. | • Percent Asymmetry (%): Ratio of the velocity of leftward versus rightward eye movement recorded during stimulation. |
| Active Head Rotation<br>(Horizontal, vertical) | Subject moves their head horizontally or vertically at specific velocities while maintaining their gaze on a fixed target. | • Gain and symmetry. Ratio of head and eye velocity |

Classification of severity were created from the Symptom Score by using the structure of the right-tailed nonparametric distribution from the normative group (Acute Group) with the method demonstrated in Lovell et al. (2006). We identified into five classifications of symptom severity (Low-Normal, Broadly-Normal, Borderline, Very High) that corresponded to the Symptom Scores cut offs at the 50^th^, 75^th^, 90^th^, and 98^th^ percentiles. Participants from the non-normative group (Long COVID Group) were placed in these classifications according to their Symptom Score expressed as a % of the group.

In addition to the concussion-related symptoms included in the symptom questionnaire, we asked participants to indicate the presence and severity following infection of three symptoms specific to SARS-CoV-2, loss of Taste and Smell, Fever, and Muscle Weakness, but not associated with Concussion. Participants were prompted to list any symptoms not included in the questionnaire that came about after infection.

### Eye Tracking with Videonystagmography

We performed an abbreviated vestibular evaluation including ocular motility assessment using infrared video-nystagmography (VNG) to assess deficits in central and peripheral sensorimotor integration. Ocular motility tests can often highlight specific areas of the brain that exhibit deficits for purposes of lesion localization. The VNG assessment consisted of five subtests. We performed oculomotor studies of smooth pursuits (tracking), randomized saccades, and optokinetic stimulation. We also examined VOR (peripheral vestibular-ocular reflex) function with active head rotations and caloric stimulation, though the latter data was not utilized for purposes of this study. Each ocular motility subtest or parameter was quantitatively compared to system normative data to determine normalcy. Additionally, for some subtests we also qualified findings based upon specific morphological features (e.g., smooth pursuit testing was qualified as normal (fluid) versus saccadic when appropriate).

For lesion localization, we examined smooth pursuits, random saccades, and OPK. The smooth pursuits parameters investigated included gain measures at 0.2 and 0.4 Hz. Symmetrically impaired pursuits have been reported in individuals with diffuse cortical, basal ganglia and/or cerebellar dysfunction. Asymmetric pursuits are often associated with focal lesions involving the ipsilateral cerebellar hemisphere, brainstem, or parieto-occipital regions. Saccade parameters investigated included saccadic velocities, latencies, and accuracy. Velocity abnormalities can be seen in individuals with basal ganglia, brainstem, cerebellar and/or lesions of the peripheral oculomotor nerves or muscles (typically diffuse lesions of central pathways commonly associated with neurodegenerative diseases). Accuracy abnormalities are typically seen with disorders of the cerebellar flocculus (hypometria) or cerebellar vermis (hypermetria). OPK eye movement is dominated by a reflexive pattern which includes both tracking (foveal) and OPK (retinal) receptor stimulation when tested with VNG.

The VNG data were read by a blinded clinician (author TH) for presence of central lesion and overall impressions. When the finding is abnormal, the categorized into Central, Peripheral, Non-localizing lesion.

### King-Devick Rapid Reading Test

*We used the King-Devick (KD) rapid number reading test, which functionally combines* saccadic eye motion and *neurocognitive demand*. The KD has shown to have a high specificity, 86% correct, for predicting concussions (Galetta et al. 2016). The subject is presented with four number reading cards that gradually increase in difficulty. The test is designed with a habituation anticipated; therefore, the difficulty of the number reading task increases as habituation occurs. The participant reads the numbers in order on each card aloud as quickly and accurately as possible and their time and errors are counted. The first card is a warmup and the last three are recorded, then the times for the three cards are summed to obtain a “Total KD.” Although it is easy to collect, the error counting hasn’t been an effective clinical measure, so we will not report number of errors in this manuscript and focus on the time taken to perform the task for each card.

### Neurocognitive Assessment with ANAM

We used a subset of the Automated Neuropsychological Assessment Metrics (ANAM) to assess neurocognitive processing and reaction time for each participant (ANAM; Reeves et al., 1995; Vista LifeSciences, Parker, CO). The ANAM battery measures a variety of neurocognitive abilities such as reaction time, emotional regulation, attention, memory, visuospatial skills, and inhibition (Reeves et al., 1995). It is a substantially validated measure (Bleiberg et al., 2000; Jones et al., 2008; Kabat et al., 2001) with strong test-retest reliability (*r* = .91; Vincent et al., 2017). The assessment was administered on a tablet device, and the battery consisted of six subtests: SRT, Simple Reaction Time; Procedural Reaction Time; MP, Mathematical Processing; M2S, Matching to Sample; SR2, Second administration of SRT; and GNG, Go-No Go. Those tests measure Simple Reaction Time, Procedural Reaction Time, Learning, Spatial Working Memory, and Inhibition (Reeves et al., 1995). We focus our analysis on the subtest Simple Reaction Time and Procedural Reaction time in this report. Simple Reaction Time is a fundamental measure of simple visual attention and ability to detect a single stimulus that appears at varying intervals while Procedural Reaction Time measures provides additional cognitive overhead during the reaction time task. The dependent variable was mean reaction time for both subtests since more complicated measures (e.g., throughput dependent variable) may obscure subtle changes in function (Tate, et al., 2013).

### Balance Performance Test

Each participant performed a series of standing balance tests to evaluate an element of performance tied directly to central integration. During balancing, the central nervous system must reconcile disagreement between visual, somatosensory, and vestibular systems. Standing balance is a somatosensory system dominant task with less involvement of the vestibular system. Center of pressure measures are while generally very sensitive to many pathology groupings but cannot specify which neural structures are responsible for the changes in motor output. For this study we administered the Clinical Test of Sensory Interaction on Balance (CTSIB). The subject stood as still as possible on a measurement board for four twenty second trials: eyes open on a hard surface, eyes closed on a hard surface, eyes open on a foam surface, and eyes closed on a foam surface. Four load cells were sampled every 40 ms, built in software passed the values through a 2^nd^ order low pass Butterworth filter with cutoff frequency set to 4Hz. The total center of pressure distance traveled was calculated, this path length is an indicator of postural control and well known to be sensitive to group differences.

### Statistical analysis

The Shapiro-Wilks test was examined as a distribution normality test (*p* > .05), and the inspection of a boxplot was used to determine the existence of outliers. Outliers were present in the data, so nonparametric tests were used for that reason. We considered any dependent variable important when the statistical significance was *p*<0.01 and the corresponding associative variable was strong (Cohen’s *d*>0.8, Cramer’s Phi or V > 0.50). To control confounding variables, we calculated the Pearson correlation coefficient between Age and each continuous dependent variable. To be conservative, we considered Age as a confounding actor when the dependent variable had a moderate with Age (*r*>0.5). Multicollinearity was assessed for all variables through inspection of the correlation coefficient. Pearson correlation coefficient was interpreted using the criteria: fair (r=0.25), moderate to good (r=0.5), and good to excellent (d=0.7) (Portney and Watkins, 2019)

#### Categorical variable analysis

A chi-square test for association was conducted between the COVID grouping (Acute COVID and Long COVID) and categorical variables. The categorical variables used in the analyses were *symptoms from the Concussion Symptom Inventory* and *clinical impressions. Symptoms from the Concussion Symptom Inventory* and *clinical impressions* were dummy coded to indicate presence or absence of the symptom and central lesion. The number in each group (as % of the group) that fell within each level of symptom severity was calculated and compared between groups. The number of participants in each group (as %) that marked a listed symptom from the post-concussion symptom questionnaire were compared across groups. All expected cell frequencies were greater than five. The Relative Risk Ratio was examined for the probability that Long COVID would occur if a pre-existing Central Lesion was present. Cramer’s Phi was calculated for each group that had a statistically significant Chi squared test for independent samples to examine the strength of association. Cramer’s V was calculated to examine the strength that the distribution across the 5 classifications. The interpreted strength of association was done using the criteria: low (phi = 0.1), moderate (Phi = 0.3), and high (Phi=0.5) (Crewson, 2016).

#### Continuous variable analysis

An independent samples t-test was performed comparing the COVID grouping (Acute COVID and Long COVID) and continuous variable scores. The continuous variables used in the analysis were *vestibular ocular* and *neurocognitive*. The *vestibular ocular* subtests used in the analysis were: saccades (velocity, accuracy, and latency), smooth pursuits (0.2Hz Gain, 0.4 Hz Gain), optokinetic (% asymmetry), and the King Devick Reading Test (trial 1, 2, 3 and trial 1-3 average). The *neurocognitive* subtests used in the analysis were simple reaction time, and procedural reaction time. The independent samples t-test was examined assuming unequal variances, and 95% confidence intervals are reported. Effect size is interpreted using the criteria: small (d=0.2), medium (d=0.5), and large (d=0.8) (Portney and Watkins, 2019).

## Results

### Sample Population Characteristics

Sixty-one people from the Denver, CO metro area participated in the investigation that occurred from March 1 through September 5, 2022, with 28 self-reporting to the Acute Group and 33 to the Long COVID group. The population was mostly female in both groups, which is common in voluntary research studies. The almost 1.6:1 ratio of females-to-males is just below the 95%CI: 1.8-6.2 reported in a key investigation on patient characteristics with Long COVID (Bai et al. 2022). An artifact of our recruiting methods was a statistically significant difference in age between the groups (t=6.89, *p*<0.01, *d*=1.71 [CI: 1.13-2.27]) despite the large and overlapping age ranges in each group. There were no statistically significant differences between groups in mass; however, the Long COVID Group was infected 9 months prior to the Acute Group (t=4.74, *p*<0.01, *d*=1.16 [CI: 0.63-1.69]) at the time of testing (Table 4).

**Table 4:**
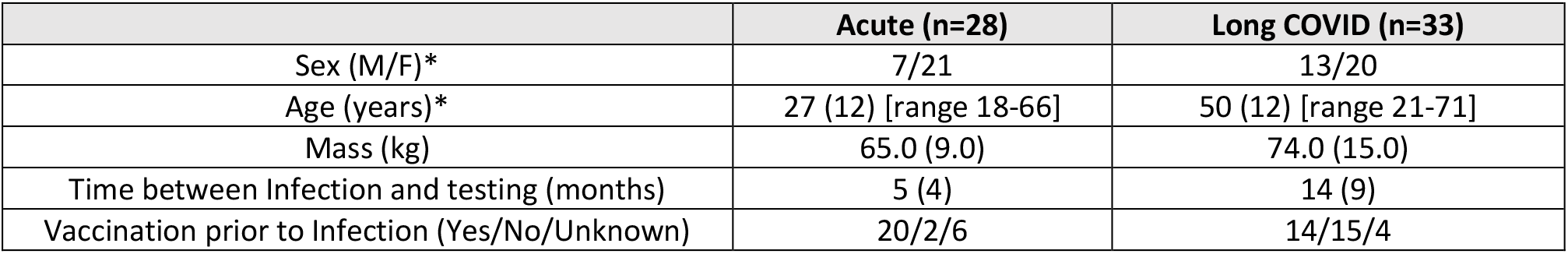
Participant characteristics. Data listed as mean (SD) or numbers with range given.* indicates statistically significant difference between the group means. Categorization into Acute and Long COVID groups was through participants self selection.

Twenty of 28 (71%) in the Acute Group were vaccinated for SARS-CoV-2 prior to infection compared to 14 of 33 (42%) of Long COVID Group. Although these ratios are statistically different (*χ*^2^=4.06, *p*=0.04), the association of vaccination status and group is weak (*φc* = 0.29). This weak association is consistent with mixed results among emerging investigations that explored the association between vaccination for COVID-19 and developing Long COVID (Notarte et al. 2022). Eight of 28 participants (29%) in the Acute Group and 17 of 33 (52%) in the Long COVID group answered *Yes* to the question “Please list any medical conditions you had prior to having COVID-19” and then provided the prior medical conditions on the similar (Table 5). These ratios were not statistically different from each other (*χ*2= 2.42, *p*=0.12) and only weakly associated (*φc* = 0.23).

**Table 5:**
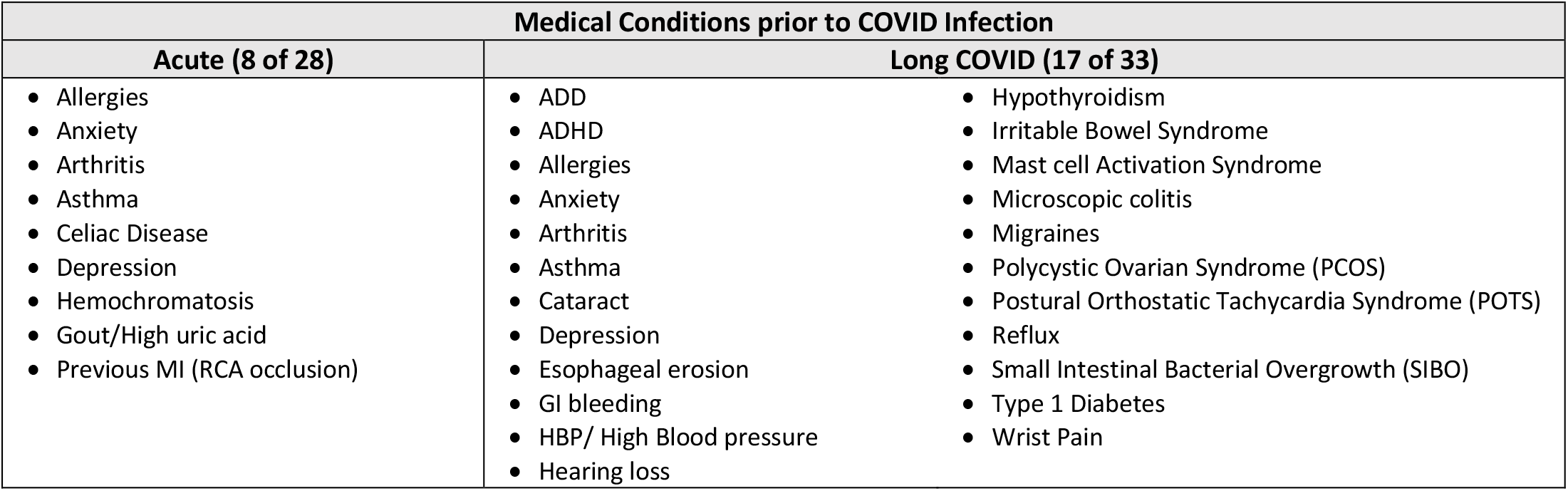
Prior medical conditions reported in response to the question “Please list any medical conditions you had prior to having COVID-19”.

### Post-concussion Symptom Questionnaire as a Sensitive Tool for Long COVID

Every one of the 22 symptoms in the post-concussion symptom questionnaire were statistically associated (Pearson *χ*^2^ squared test, *p*<0.05) with the self-reported presence of Long COVID, and 21 of those were highly statistically significant (p<0.01) (Table 6). Presence of the nine Somatic Symptoms varied among the Long COVID group while cognitive, sleep, and affective symptoms were present in almost all participants in the Long COVID group. The most common Somatic symptoms reported in the Long COVID group were Headache (1 out of every 2), Balance Problems (4 out of every 5), Dizziness (3 out of every 4), and Blurry or double vision (1 out of every 2). Each of these somatic symptoms were highly associated with Long COVID except Headache. The Low association of Headache is the result of many Acute participants (1 out of 5) also reporting the onset of headaches following COVID infection. Except for headache, the Acute group had very low prevalence of somatic symptoms.

**Table 6:** Percentage of each group (Acute and Long COVID) that reported each symptom. Double asterisks indicate that the statistical significance is *p*<0.01 and the symptom has moderate (***φ***_***c***_ > 0.4) or high association (***φ***_***c***_ > 0.5) with the Long COVID group. The last three symptoms (listed in the gray box) were not part of the post-concussion symptom questionnaire.

| Symptoms and Category | Acute (n=28) | Long COVID (n=33) | Pearson's $\chi^2$ | Cramer's $\phi_c$ |
| --- | --- | --- | --- | --- |
| **Physical/Somatic Symptoms** |  |  |  |  |
| Headache** | <u>18.8%</u> | <u>54.5%</u> | $\chi^2 = 8.94, p < 0.01$ | Low: 0.37 |
| Nausea** | 3.1% | 39.4% | $\chi^2 = 12.64, p < 0.01$ | Moderate: 0.44 |
| Vomiting* | 0.0% | 12.1% | $\chi^2 = 4.13, p = 0.04$ | Low: 0.25 |
| Balance Problems** | 3.1% | <u>78.8%</u> | $\chi^2 = 38.3, p < 0.01$ | High: 0.77 |
| Dizziness** | 0.0% | <u>72.7%</u> | $\chi^2 = 36.90, p < 0.01$ | High: 0.75 |
| Blurry or double vision** | 3.1% | <u>54.5%</u> | $\chi^2 = 20.77, p < 0.01$ | High: 0.56 |
| Sensitivity to Light** | 6.3% | 45.5% | $\chi^2 = 12.93, p < 0.01$ | Moderate: 0.45 |
| Sensitivity to Noise** | 9.4% | 48.5% | $\chi^2 = 12.01, p < 0.01$ | Moderate: 0.43 |
| Pain other than Headache** | 3.1% | 42.4% | $\chi^2 = 14.14, p < 0.01$ | Moderate: 0.46 |
| **Cognitive Symptoms** |  |  |  |  |
| Feeling “in a fog” ** | <u>21.9%</u> | <u>93.9%</u> | $\chi^2 = 34.74, p < 0.01$ | High: 0.73 |
| Feeling Slowed Down** | <u>21.9%</u> | <u>100.0%</u> | $\chi^2 = 41.90, p < 0.01$ | High: 0.80 |
| Difficulty Concentrating** | <u>25.0%</u> | <u>97.0%</u> | $\chi^2 = 35.55, p < 0.01$ | High: 0.74 |
| Difficulty Remembering** | <u>25.0%</u> | <u>93.9%</u> | $\chi^2 = 32.17, p < 0.01$ | High: 0.70 |
| **Sleep Symptoms** |  |  |  |  |
| Trouble Falling Asleep** | 28.1% | <u>63.6%</u> | $\chi^2 = 8.24, p < 0.01$ | Low: 0.36 |
| Fatigue or low energy** | 34.4% | <u>97.0%</u> | $\chi^2 = 28.42, p < 0.01$ | High: 0.66 |
| Drowsiness** | 18.8% | <u>75.8%</u> | $\chi^2 = 21.16, p < 0.01$ | High: 0.57 |
| **Affective Symptoms** |  |  |  |  |
| Feeling more Emotional** | <u>25.0%</u> | <u>75.8%</u> | $\chi^2 = 16.75, p < 0.01$ | High: 0.51 |
| Irritability** | <u>21.9%</u> | <u>69.7%</u> | $\chi^2 = 14.95, p < 0.01$ | Moderate: 0.48 |
| Nervousness** | 12.5% | <u>60.6%</u> | $\chi^2 = 16.14, p < 0.01$ | High: 0.50 |
| Sadness** | 12.5% | <u>60.6%</u> | $\chi^2 = 16.14, p < 0.01$ | High: 0.50 |

| **Additions to Post-Concussion Symptom Questionnaire** |  |  |  |  |
| --- | --- | --- | --- | --- |
| Loss of Taste or Smell** | 9.4% | 42.4% | $\chi^2=9.19, p<0.01$ | Moderate: 0.38 |
| Fever | 3.1% | 9.2% | $\chi^2=2.08, p=0.09$ | — |
| Muscle Weakness** | 6.3% | 41.5% | $\chi^2=32.32, p<0.01$ | High: 0.71 |

All four cognitive symptoms (Feeling “in a fog”, Feeling Slowed Down, Difficulty Concentrating, Difficulty Remembering) were highly associated with Long COVID and a notably high proportion of the Acute Group also reported new onset of cognitive symptoms following COVID infection. It is notable that almost 100% of the participants in the Long COVID reported the presence of all the Cognitive symptoms. The Acute Group also reported onset of these Cognitive symptoms at approximate rates ranging from 20-25%.

Sleep Symptoms demonstrated a similar pattern to the Cognitive Symptoms with a high proportion of the Long COVID group experiencing the symptoms (e.g., almost 100% for the symptom of Low Energy) with approximate rates in the Acute group of 20-35%. All four of the affective symptoms had medium or high level of association with Long COVID. Feeling more emotional, nervousness, sadness had high associations, and association of Irritability was close to a high association. Rages for the Long COVID participants to report affective symptoms were approximately 2 out of every 3. It is notable that participants in the Acute Group also reported non-zero rates of Feeling more Emotional (1 out of 4) and Irritability (1 out of 5) as an onset following COVID 19 infection.

### Further Long COVID distinguishers using Additional Symptoms

Of the three non-concussion-related symptoms we added to Post-Concussion Symptoms Questionnaire, Muscle Loss of Taste and Smell and Muscle Weakness were statistically significant, and had moderate and high association with Long COVID, respectively. Despite listing “intermittent fever” by the WHO as a common Long COVID symptom, fever had the fewest participants in the Long COVID group report this as a symptom (9.2%). When invited to list any additional symptoms that the participant since infection 18 out of the 33 in the Long COVID Group reported symptoms in while only 2 out of the 28 in the Acute Group reported symptoms. Many of the additional symptoms reported by the Long COVID group are be placed in the Autonomic, Neuroendocrine, and Immune symptom categories (Table 7), which were not included in the post-concussion symptom questionnaire. Additional symptoms were also placed in the Somatic and Sleep categories. No additional symptoms were added to the Cognitive or Affective symptom categories, which indicates the Post-Concussion Questionnaire sufficiently covered those categories. Three participants in the Acute Group listed additional symptoms of Rash, Pericarditis, and Cough.

**Table 7:**
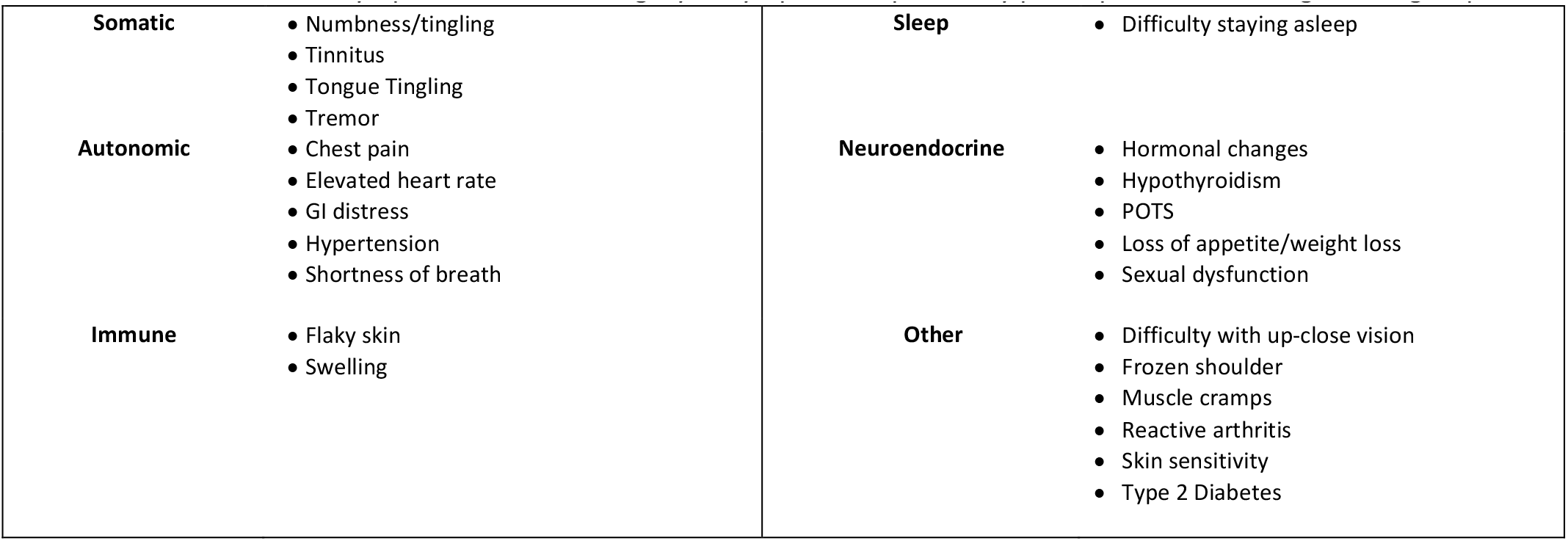
List of additional symptoms and the category of symptoms reported by participants in the Long COVID group.

### Severity of Long COVID from the Post-concussion Symptom Questionnaire

Participants from the Acute Group reported few symptoms, and which resulted in a Total Symptom Scores that fell at the low end of the possible range of scores, and a severely right-tailed distribution (Shapiro-Wilk’s *W*=0.726, *p*<0.01) similar to the previous analyses of healthy participants in the Post-Concussion Scale (Lovell et al., 2006). By contrast, Total Symptom Scores from the Long COVID group spanned almost the entire range of possible scores and created a more normal distribution (Shapiro-Wilk’s *W*=0.973, *p*=0.55).

The 50^th^, 75^th^, 90^th^, and 98^th^ quantiles natural distribution of the Acute Group created classification where the with a Normal range (Low-Normal + Broadly-Normal) of 0–11, a narrow Borderline Range of 12–18, and very large High Range of 19–128 (Figure 1). When participants from the Long COVID group were classified using this scheme, only 2 of 33 (6%) were classified in the Normal range, 3 of 33 (9%) were classified in the Borderline range, and 28 of 33 in the High Range. The five-category classification of the participants were statistically different across the Acute and Long COVID groups (*χ*2= 39.44, *p*<0.01) and the distribution across the classifications was highly associated with the group (Cramer’s *V* = 0.80).

**Figure 1.**
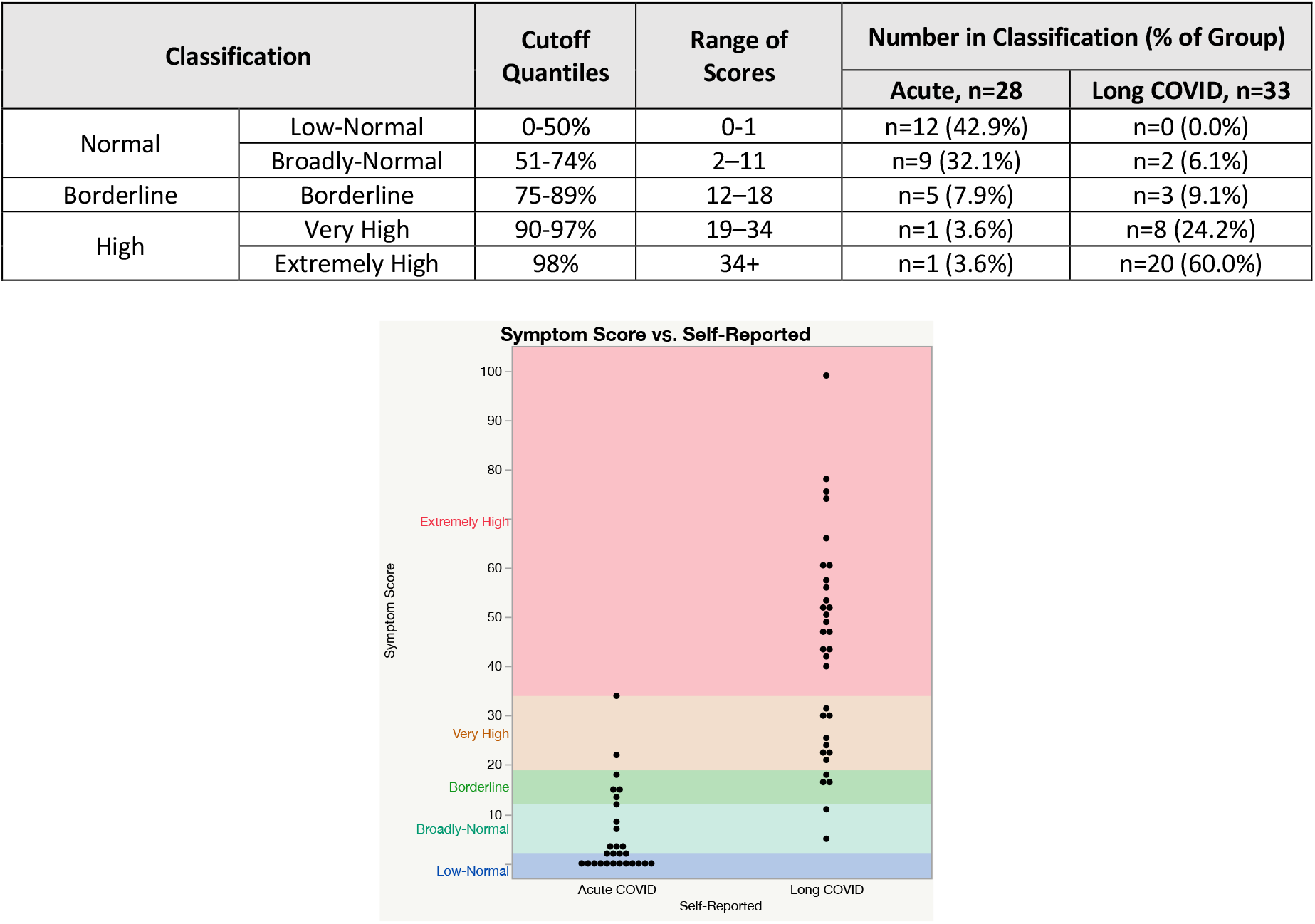
Percentage of each group (Acute and Long COVID) classified in five levels based on the distribution of a normative sample population (Acute COVID).

### Presence of Central Lesions observed within VNG Recording

Only 1 of the 28 (3.6%) participants from the Acute Group demonstrated signs of a central lesion, while 5 of the 33 participants (15.1%) from the Long COVID group and indicates the presence of a central lesion. All clinical findings of central deficits in the Long COVID group occurred during the random saccade subtests. Three participants from the Long COVID group demonstrated reduced saccadic velocities, which indicates the presence of lesions in the basal ganglia, brainstem, or cerebellum (Table 8. Two participants from the Long COVID group demonstrated hypermetric saccades, which indicate a lesion within the cerebellar vermis. One of the 28 participants from the Acute group demonstrated signs of a central lesion, which consisted of saccadic intrusion during the Smooth Pursuits test (Table 8). Although the presence of central deficits was different from zero in the Long COVID Group (95% CI: 6.7–30.9%), when were compared to the Acute group there was no statistical difference (χ^2^=2.29, *p*=0.13, *φc*=0.13). If we assume that the central lesions detected from the VNG over-reads existed prior to being infected by COVID, the presence of the lesion increases the risk of developing long COVID by 64% (Relative Risk=1.64 [CI: 1.05–2.54]).

**Table 8:**
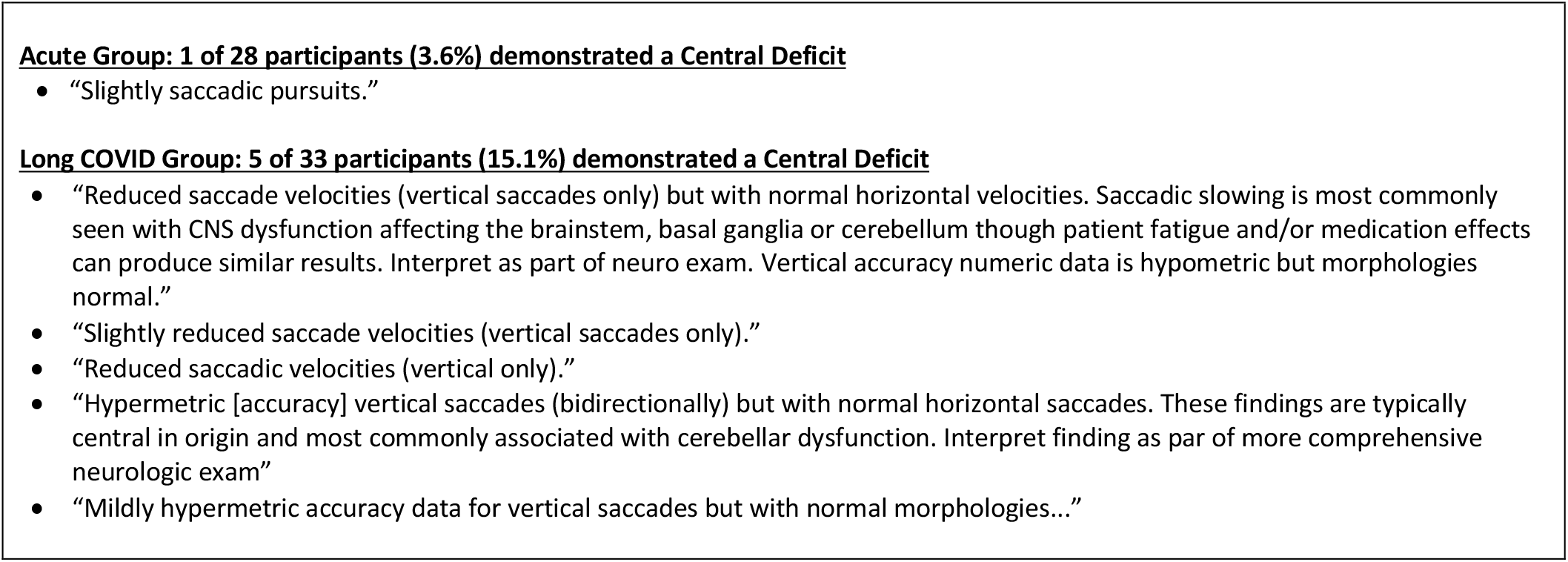
Clinical impressions from blinded VNG Overreads performed by the author (TH). Five participants (of 33) in the Long COVID group demonstrated a Central Deficit while only

### Limited Sensitivity of Quantitative VNG Subtests

Saccadic latency measured during the Horizontal Random Saccades was the only variable within the quantitative VNG subtests that was statistically significant to the effect of group and with a large effect size. Saccadic latencies in the Long COVID Group were 22 msec longer on average than in the Acute group (Figure 2). Normal latency is approximately 200-220 msec (Hopf et al., 2018). Range of the data in the Acute Group bracketed this standard. By contrast, most participants in the Long COVID group exceeded this standard, with data ranging from 205 to 309 msec. Highest 15% of the Long COVID had average latencies over 280 msec. Saccadic latency can be higher in patients with neurodegenerative diseases (Hopf et al., 2018; Antoniades and Kennard 2015) and other aging processes. However, the low correlation of age and horizontal saccadic latency (*r*=0.47) indicates that Age was not a confounding factor in this outcome. The similar effect of Long COVID on latency was not present in the vertical saccades. Dependent variables from the Smooth Pursuits and Optokinetic subtests were not sensitive to group differences.

**Figure 2.**
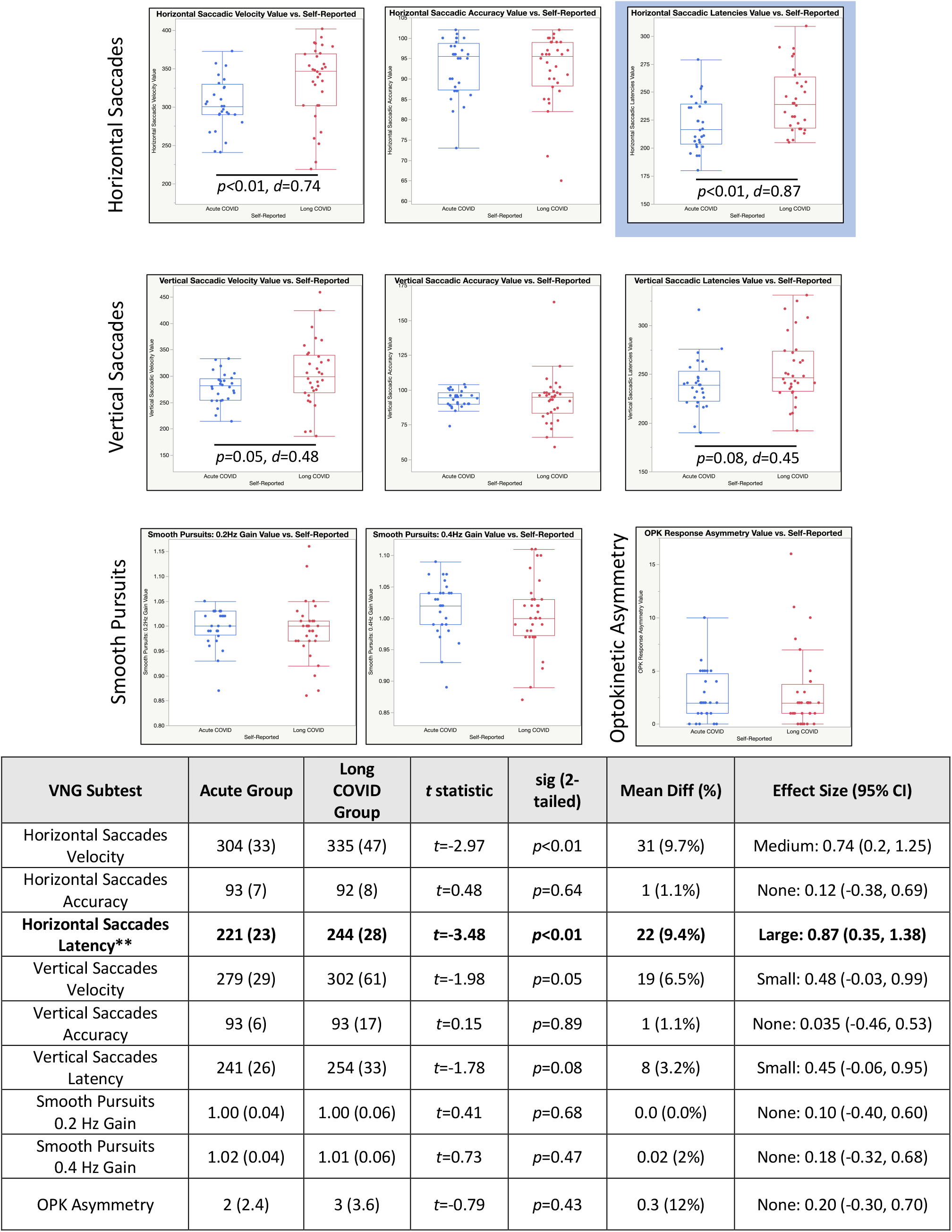
Results from the battery of video nystagmography (VNG) eye tracking subtests. Saccadic latencies (in msec) with recorded during in the Horizontal Saccades subtest was statistically significant with a Large effect size. Saccadic latencies in the vertical direction was also statistically significant, but demonstrated a Medium effect size.

The time taken to read in all three King-Devick Cards and the Total KD composite of the three cards was higher in the Long COVID Group compared to the Acute Group (Figure 3). All four dependent variables were highly statistically significant (p<0.01) and had large effect sizes that ranged from 1.00 to 1.47. Age was not correlated with any of the four dependent variables (r=0.27, 0.22, 0.21, 0.24), so is not considered a confounder in the KD. The difference in means of 19.2 sec is much larger than the standard error of measurement, SEM=2.29 sec, and over 3 times larger than the minimum detectable change, MDC=6.35 sec (Heick, 2016).

**Figure 3.**
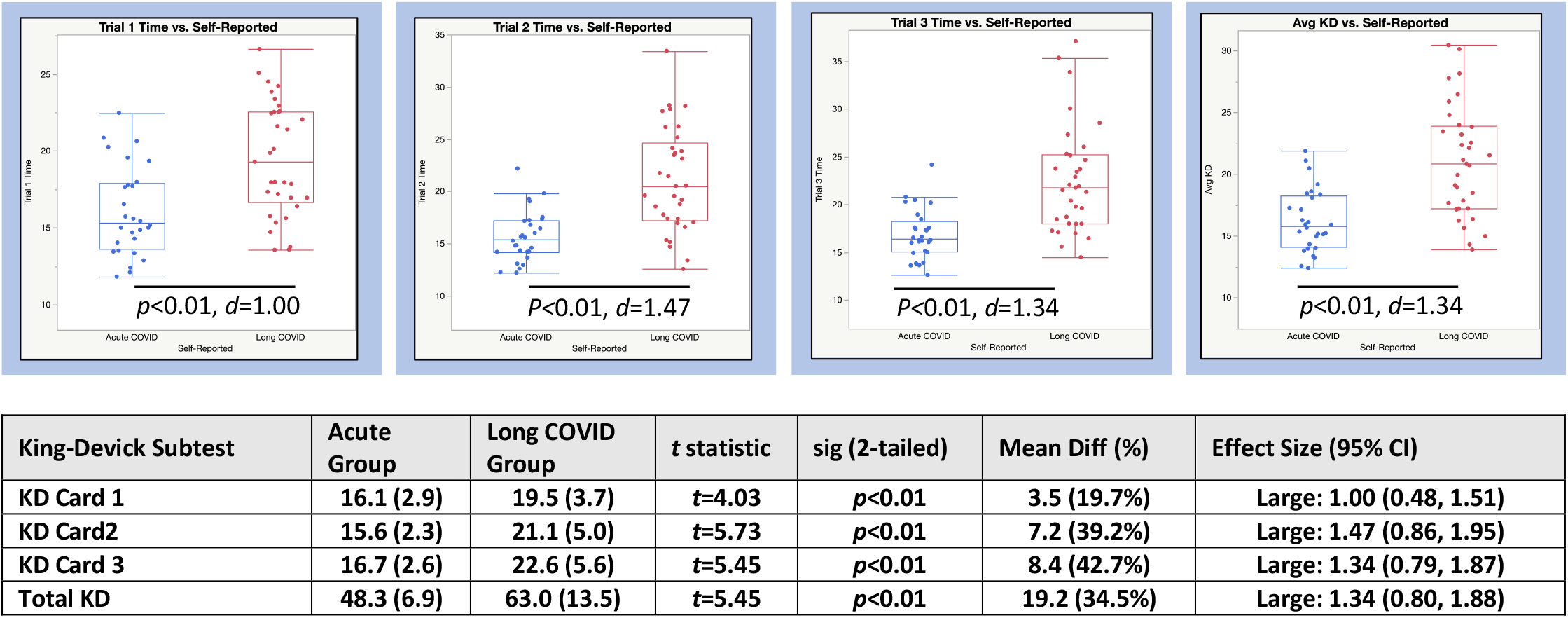
Time taken (in sec) for all three trials of the King-Devick reading test and the Total KD (sum of the three trials) were highly sensitive to the effect of Long COVID with statistically significant differences between groups with Large effect sizes.

### Reaction Time Sensitivity in Subtests from the Automated Neuropsychological Assessment Metrics (ANAM)

The Long COVID Group was slower than the Acute Group on the Simple Reaction Time and Procedural Reaction Time subtests of the ANAM (Figure 4). The differences were highly statistically significant (*p*<0.01) and the effect sizes on each test were large (*d*=0.93 and *d*=1.09, respectively). Age was moderately correlated with Simple Reaction Time (*r*=0.53) and which indicates some confounding of Age on the effect of Group in this variable, and only fairly correlated with Procedural Reaction Time (*r*=0.41).

**Figure 4.**
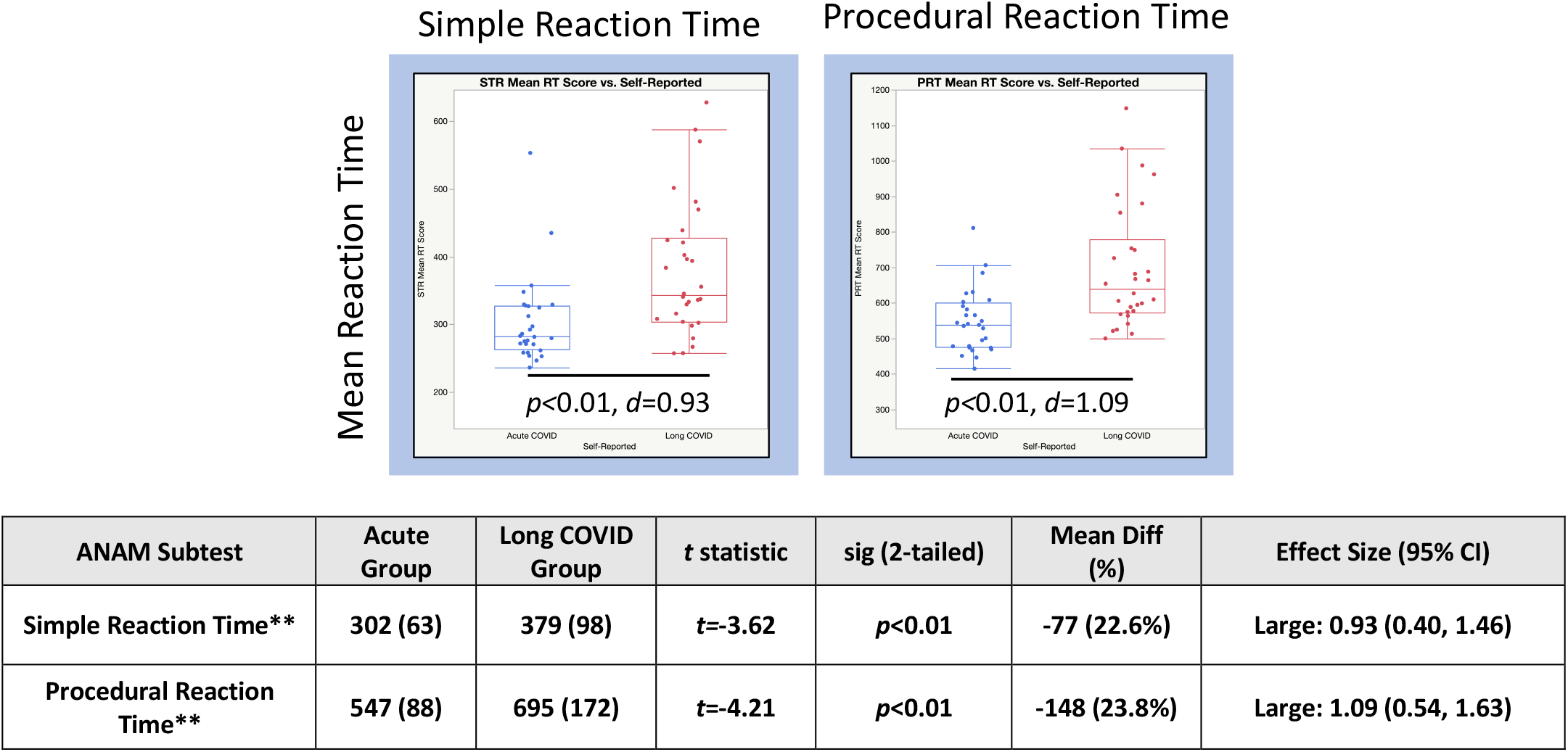

### Sensitivity in Balance Performance Subtests

The Long COVID Group had a longer center of pressure pathlength than the Acute Group on all subtests in the balance battery. Each subtest that involved a form of sensory deprivation (e.g., Eyes Closed, Soft surface), were statistically significant to group with Medium to Large effect sizes (Figure 5). Age was moderately correlated with the subtest that combined both Eyes Closed and Soft Surface (*r*=0.60), which indicated some level of confounding by Age on the group effect. The other statistically significant subtests, Eyes Closed Hard Surface and Eyes Open Soft Surface, demonstrated fair correlations (r=0.32 and r=0.37, respectively). The difference in pathlength during a 20 second collection of 7.0 to 9.5 cm in the Eyes Closed Hard Surface and Eyes Open Soft Surface are larger/smaller than the standard error of measurement, SEM=### msec, and smaller/larger than the minimum detectable change, MDC=### msec,(ref). Add sentence on how the mean difference compares with Moira’s paper on acute concussion.

**Figure 5.**
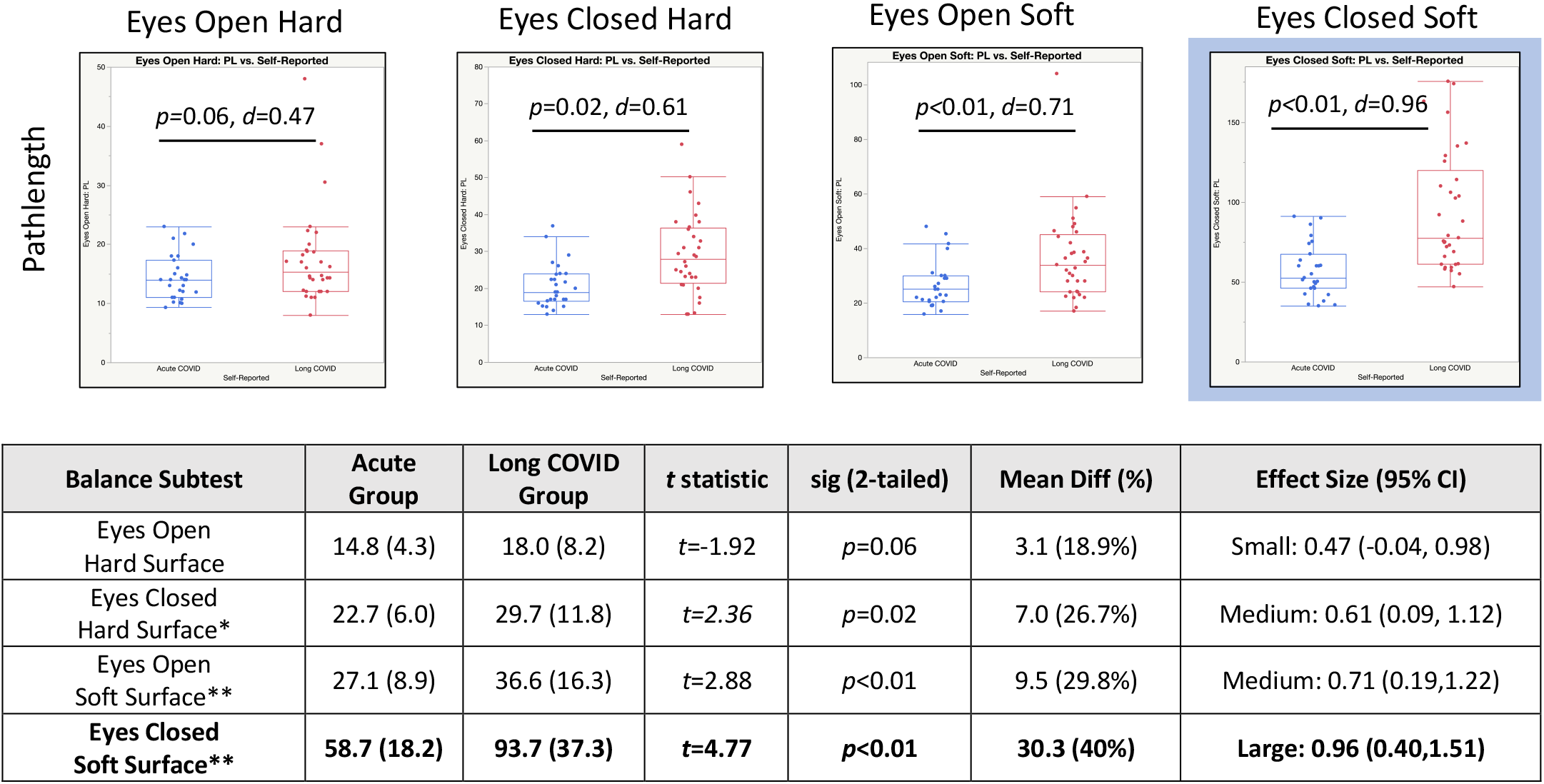
Mean pathlength (in cm) for each subtest of the balance performance assessment. * indicates *p*<0.05 and ** indicates *p*<0.01. Dependent variables with statistically significant differences (*α*=0.05) and Large effect sizes shown in bold.

## Discussion

In this investigation, we evaluated whether instruments of post-concussion evaluation could be used to differentiate between participants with Long COVID and a reference group who only experienced an Acute infection. Verifying that a post-concussion framework can be used in a Long COVID patient population opens new avenues of diagnoses and treatment for a condition that is currently affecting millions of patients and has proven challenging to treat. Our results indicate that long-standing and validated post-concussion symptom questionnaires may be used for quantifying the severity of Long COVID. A wide variety of objective and quantitative measures from post-concussion care are sensitive to the Long COVID condition. Some of the most sensitive measures—especially the King-Devick rapid reading test—may be effective at tracking patient progress in the context a Long COVID treatment. Although central lesions were not a prominent feature in this Long COVID population, the results point to wide deficits in motor integration. These results also provide a rationale for treating the subset of Long COVID patients with clear neurological symptoms with similar rehabilitation strategies as patients with post-concussion syndrome. As treatments emerge for patients suffering from long COVID, the accurate and objective measurement of patient progress (e.g., symptom reduction and functional performance) will be critical.

Post-concussion symptom questionnaires with a wide range of symptoms and a large scoring range can be used as a starting point for evaluating severity of Long COVID symptoms and could be a tool for judging the effectiveness of therapeutics. A key feature of the post-concussion instrument is that it includes a clear affective category which doesn’t exist in other disease models such as ME/CFS. Using the post-concussion symptom questionnaires, very few self-reported Long COVID participants were classified into a normal self-reported symptom range. A small number of re-classifications may be expected given that this scale is a “state measure of perceived symptoms on a particular day”. Current data from this particular tool indicates that patients who score above 12 could use additional symptom evaluation for Long COVID.

Because tools built on concussion do not encompass the full range of symptoms possible, direct questioning on other common symptoms of Long COVID should be employed. This highlights a limitation of the instrument, which is that it does not include neuroendocrine, autonomic, and immune function questions. In addition, we have a limited sample size, and it would be interesting to examine how these results stand up in larger populations. Regardless, we believe the post-concussion assessment provides a good starting point. There exists multiple options for symptom questionnaires with a wide range of language, resolution in quantification, and descriptions. We prefer a tool with a large range of symptoms and high resolution so that a measure is as continuous as possible, like the Continuous-Scale Physical Functional Performance test (CSPFP) in functional movement assessments in older adults (Cress et al. 2005).

The post-concussion symptom questionnaire may also be effective to identify patients who are not classified as Long COVID but could benefit from therapy. For instance, a notable number of participants in the Acute group indicated symptoms and could possibly benefit from treatment. One out of four Acute participants reported brain fog, fatigue, and affective symptoms, which are both typical of Long COVID. These symptoms, even at low levels, can adversely affect daily performance. Given that the symptom questionnaires measure the current “state” (Lovell et al., 2006), it is likely that reductions in the Total Symptom Score can be achieved in a borderline case with the right therapeutic approach.

The inventory can be substituted with any common inventory such as the Concussion Symptom Inventory (Randolph et al., 2009), SCAT-5 Inventory (Echemendia et al. 2017), or the Post-Concussion Symptom Inventory (Gioia et al., 2008). If symptoms are used as an intervention target then a high resolution without a ceiling effect would be advisable. Studies indicate that Long COVID primarily has neurological origins and therefore, may be considered a neurological disease process; post-concussion syndrome and ME/CFS are also categorized as neurological disorders. Therefore, any symptom inventory employed should focus in this area.

Tests that require rapid eye movement suggest that voluntary saccadic motion can be used in clinical and research-based evaluation of Long COVID. In the current study, latency in random saccade tests were the most sensitive of the quantitative VNG measurements. Although these values did not indicate an acute lesion (see previous section on clinical findings from over-reads), the prolonged latencies compared to the Acute group suggest slower central processing related to pathways in the frontal cortex or basal ganglia. Saccadic latency is an important functional component of planning and execution of whole-body movement and fine motor tasks such as reading (see next paragraph). Prolonged latency can affect a person’s ability to respond effectively to motor situations like driving or playing sports, etc. The inability to move the eyes with precision can lead to headaches during reading and other focused tasks. Further, a delay in reaction time is likely to be perceived as mental fogginess or sluggishness, two of the most common complaints among persons with Long COVID. Our findings are consistent with Douaud et al. (2022) that reported a significantly greater increase in the time taken to complete a trail-making test among the participants with SARS-CoV-2 infection. Predictive saccades and anti-saccade tests, which require both reflex inhibition and saccades to be generated volitionally, may be more effective than random reflexive paradigms in which the participant need only follow a target. Future research should use tests with pro- and anti-saccades of the demand for executive function.

Of the tests we performed, the King-Devick incorporates the most elements affected by neurological deficits and is highly functional and therefore, may have the highest usefulness in the clinic among the tests examined. The KD rapid number reading requires precise saccadic motion and demonstrated the highest sensitivity of all the dependent variables in the study. Procedural reaction time also incorporates elements of executive function for a localized motor control task and was very sensitive to Long COVID. This follows from the success that it has had of predicting concussion (Galetta et al. 2016). A notable limitation here is that our study used baseline data on all the participants for comparison. In addition, there can be no absolute diagnostic cutoffs since a “perfect score” doesn’t exist, like in the Total Symptom Score. Nonetheless, the fact that the change in KD was sensitive, this indicates that the Total KD could be used as a tool to examine the effect of therapy by administering it before and after therapy. To our knowledge this hasn’t been performed. Because spontaneous healing in the vast majority of concussion patients occurs rapidly— typically within two weeks—the KD doesn’t make for a timely measure to track healing. By contrast, because most people suffer from Long COVID in a chronic sense with little if any spontaneous healing, using the KD may provide a timely and sensitive test to examine the effects of Long COVID treatment, regardless of the treatment applied (e.g., physical therapy, cognitive training, neurological pharmaceuticals, etc.).

The assessments of neurocognitive performance and ability to maintain balance can add valuable information about how central integration deficits from Long COVID affect daily function. In this study, the neurocognitive and balance measures were very sensitive to group differences. Specifically, the long COVID group’s performance reflects balance problems including dizziness and poorer neurocognitive functioning like slower processing speed and reaction time. That delay in reaction time is likely to be perceived as mental fogginess or sluggishness, two of the most common complaints reported by persons with Long COVID. This slowing of motor and processing speed is consistent with Douaud et al. (2022) who reported a significantly greater increase in the time taken to complete a trail-making test among study participants with SARS-CoV-2 infection. Processing speed or procedural reaction time, incorporates elements of executive function for a localized motor control task and was very sensitive to Long COVID. The balance measures also reflected deficits in motor integration. A recent study reported that balance deficits were characteristic of a young population who have had COVID 19 (Guzik et al., 2022). the Balance Effect size was greatest when all sensory information was disrupted except the vestibular system, which forced reliance on the vestibular system. This result needs further investigation however, given the confounding effect of age in this subtest.

### The current diagnostics for Long COVID remain limited, without standardization, and may benefit from longstanding instruments found in post-concussion assessment that are easy to use, quantitative, and require no capital investment

These instruments can immediately add sensitive symptom questionnaires to classify severity of illness.

Post-concussion questionnaires do a good job of focusing on the somatic, neurological, and affective symptoms. We recommend adding broad symptoms in the neuroendocrine, autonomic, and immune categories to adequately cover Long COVID patients with non-neurological subtypes. Similarly, the King-Devick can be used immediately as a functional test. It is likely that tests like the King-Devick would go largely unnoticed to the treating physicians. Computerized neurocognitive tests have a long history of use in sports, for example, to quantify the effect of concussion. Some computerized neuropsychological tests are designed to be simple to administer and assess gross cognitive and motor function. Those tests often require an investment in licensing and interpretation. Adding sensitive measures of balance provides a global view of central nervous system performance but may require an even more significant investment.

### Future applications

There is still much to accomplish in the identification and management of patients with Long COVID. We are hopeful that the long-established framework of concussion may provide readily available tools and approaches to understanding Long COVID. As a medical community, we must do well at treating the current patients and keep an eye on the long-term outlook of Long COVID. This aspect is modeled well in the concussion community and TBI, which continues to improve athlete health while also focusing on understanding and pre-empting the risk of long-term neurodegenerative disease associated with multiple brain injuries. Very recent research also reflects the relationship between COVID and brain injury where the risk for sustaining a brain injury is higher among COVID patients (Xu et al. 2022). This may be a real possibility if the overlap of Long COVID and concussion persists as we learn more about both conditions.

## Data Availability

All data produced in the present study are available upon reasonable request to the authors.

